# Early Identification of Disease Promoters of Cognitive Decline Using Inflammatory, Immunologic and Cognitive Mapping (IICM™)

**DOI:** 10.1101/2025.10.14.25337174

**Authors:** Marko Lukic, Xiao-ke Gao, Marvin Collin, Rami Cohen, John Brooks, Rohit Kulkarni

## Abstract

Dementia, including Alzheimer’s disease, is a growing global health crisis that remains underdiagnosed, with limited treatment options available once symptoms emerge. Inflammation, vascular dysfunction, and metabolic dysregulation have been identified as key systemic promoters of cognitive decline, yet they are rarely integrated into current diagnostic workflows. We present the Inflammatory, Immunologic, and Cognitive Mapping (IICM™) platform. This novel framework combines cytokine biomarker profiling, digital neurocognitive assessment, and systemic comorbidity mapping to identify root causes of decline at early and preclinical stages. In a prospective observational pilot study of 45 older adults, we developed two condition-specific indices: the Biomarker Risk Score (BRS) and Cognitive Risk Score (CRS), and two global indices of disease burden, the Global Patient Risk (GPR) and Global Cognitive Risk (GCR). Preliminary findings demonstrate the platform’s ability to detect early cognitive underperformance, correlate biomarker dysregulation with cognitive deficits, and identify modifiable risk factors such as pre-diabetes. Importantly, we also identified a “golden hour” cohort with elevated biomarker activity but preserved cognition, representing a critical window for preventive intervention. These results highlight the potential of IICM™ to enable earlier diagnosis, guide personalized risk stratification, and inform precision-based strategies for dementia prevention and care.

## Introduction

Dementia, including Alzheimer’s disease (AD) and related neurodegenerative disorders, is being increasingly recognized, highly prevalent, and severely underdiagnosed. In the U.S., more than 92% of expected cases of mild cognitive impairment (MCI) remain undocumented in Medicare claims, and nearly 80% of patients are diagnosed only at moderate-to-severe stages of dementia.^1,2^ Diagnostic rates are even lower in Black and Hispanic populations compared with non-Hispanic whites, highlighting significant disparities in care.^3^ This diagnostic gap delays access to disease-modifying therapies, care planning, and preventive interventions, perpetuating the “silent epidemic” of dementia.

The global scale of dementia is striking. In 2015, an estimated 46 million people were living with dementia worldwide, projected to exceed 130 million by 2050.^4,5^ In the United States, approximately 6.9 million older adults currently have AD, and prevalence is expected to nearly triple by mid-century.^6^ One in nine Americans ≥65 years and older already has Alzheimer’s dementia. Women constitute two-thirds of cases, while African Americans and Hispanics face up to twice the risk compared to whites.^3^ AD is now the sixth leading cause of death in the U.S., with mortality increasing by more than 145% since 2000, in contrast to declines in cardiovascular disease and cancer.^6^

Despite the scale of the problem, there are still no curative therapies. FDA-approved symptomatic agents offer temporary relief without altering disease progression. Recent monoclonal antibodies such as lecanemab and donanemab provide modest slowing of cognitive decline but are limited to early disease stages and require early diagnosis for eligibility.^7^ For many patients diagnosed late, treatment options remain palliative.

Mounting evidence implicates inflammation, vascular dysfunction, and metabolic dysregulation as central promoters of cognitive decline.^8-14^ Systemic inflammation, reflected in elevated C-reactive protein, interleukin-6 (IL-6), and tumor necrosis factor-α (TNF-α), has been linked to faster memory decline, hippocampal atrophy, and poor executive performance.^14^ In the present study, we operationalize biomarkers specifically as cytokine expression patterns to capture the systemic inflammatory burden that contributes to early cognitive decline.

Notably, in the brain, persistent activation of microglia and astrocytes sustains cytokine cascades that exacerbate β-amyloid deposition, tau hyperphosphorylation, and synaptic injury.^15^ Similarly, vascular conditions such as hypertension, atherosclerosis, and small-vessel disease contribute to ischemic injury and white matter degeneration.^16^ In contrast, metabolic disorders such as diabetes and obesity accelerate neurodegenerative pathways via insulin resistance, oxidative stress, and cytokine activation.^17-20^ Together, these systemic and CNS factors create a pro-inflammatory, neurotoxic milieu that accelerates cognitive impairment.

The economic and societal burden is immense. In 2022, U.S. healthcare expenditures for AD and other dementias reached $321 billion, projected to exceed $1 trillion by 2050.^21,22^ Medicare beneficiaries with dementia incur three times higher costs than their peers, and Medicaid payments are more than 20-fold higher. Beyond direct costs, dementia care relies heavily on informal caregivers. More than 11 million Americans provide unpaid care, totaling 18 billion hours annually, valued at $339 billion.^23^ Caregiver stress, burnout, and workforce withdrawal add further indirect losses, making dementia one of the costliest and most disruptive health conditions worldwide.

We developed the *Inflammatory, Immunologic, and Cognitive Mapping (IICM*^*TM*^) platform, which triangulates biomarker expression, cognitive test performance, and systemic comorbidities to provide a root-cause analysis of the active promoters of cognitive decline. By integrating these domains, IICM™ is designed to identify mechanistic disease promoters well in advance of irreversible neuronal damage, guiding both early diagnosis and therapeutic targeting.

The underdiagnosis and massive societal burden of dementia, coupled with the paucity of curative options, underscore the urgency of new paradigms for early detection. By mapping inflammatory, immunologic, and cognitive interactions, IICM™ represents a novel and scientifically grounded approach to uncover the biological promoters of cognitive decline and to enable individual precision strategies that may ultimately alter the trajectory of Alzheimer’s disease and related dementias.

## Methodology

We conducted a prospective observational study at a neurology clinic in New York City, NY, USA, under the supervision of Judy Gao MD PhD, a board-certified neurologist. The protocol was reviewed and approved by the Institutional Review Board (IRB# 2023-0464, February 2024), and all participants provided written informed consent before participation. All procedures complied with principles of US HIPAA Regulations and Standards.

Forty-five patients undergoing evaluation for suspected cognitive impairment were enrolled. Inclusion criteria encompassed male and female individuals aged 65 years or older, referred for memory or cognitive complaints, or presenting with possible early stages of cognitive decline. Exclusion criteria included advanced stages of mental decline that precluded participation in the Creyos™ online brain test and the inability to provide informed consent.

Peripheral venous blood (5 mL) was collected under standardized conditions. Samples were centrifuged to extract serum, which was immediately stored at –80 °C in cryovials until analysis. Serum aliquots were subsequently reconstituted and tested using a 14-cytokine panel at a CLIA-registered facility (SBH Diagnostics). The assay was optimized for sensitivity and reproducibility, ensuring reliable detection of cytokine dysregulation across medical conditions.

Cognitive performance was assessed using the Creyos™ digital neurocognitive platform, a validated tool covering multiple cognitive domains. The assessment included six standardized testing areas: episodic memory (Paired Associates), verbal reasoning (Grammatical Reasoning), mental rotation (Rotations), visuospatial working memory (Number Ladder), attention (Feature Match), and deductive reasoning (Odd One Out). Together, these tasks provided a comprehensive evaluation of executive function, working memory, reasoning, and processing speed. The results enabled the identification of underperforming brain regions, each of which was mapped to one or more medical conditions (MCs) known to affect cognition.

The Inflammatory, Immunologic and Cognitive Mapping (IICM™) platform was developed to integrate multidimensional patient data: cytokine biomarker expression, cognitive performance, and systemic comorbidities into composite indices of dementia risk. This approach enables both condition-specific profiling and global quantification of disease promoters.

### A) Biomarker Risk Score (BRS)

The BRS quantified cytokine dysregulation attributable to each medical condition (MC). For a given MC, the number of overexpressed cytokines (CKs) was divided by the maximum number of cytokines assessed for that condition, generating a normalized value between 0 and 1.

- **Example:** If 2 out of 4 cytokines linked to a condition were elevated, the BRS = 0.5.

This score reflects the relative burden of systemic inflammatory activation associated with each MC.

### B) Cognitive Risk Score (CRS)

The CRS represented the cognitive burden imposed by each MC. Using Creyos™ performance data, the number of underperforming brain regions associated with a condition was divided by the total number of areas tested for that condition.

- **Example:** If 3 out of 10 brain regions relevant to a systemic condition were impaired, the CRS = 0.3.

Thus, CRS values capture the extent to which functional cognitive domains are compromised in relation to specific systemic or neurological conditions.

### C) Condition-Level Display

Each MC was reported with two complementary parameters:

- **BRS** – extent of biomarker dysregulation.
- **CRS** – extent of cognitive dysfunction.

This dual reporting enables root-cause mapping by linking systemic inflammatory activity to domain-specific cognitive impairment.

### D) Global Patient Risk (GPR)

The GPR provides a **summative index of biomarker dysregulation** across all MCs. It was calculated as the ratio between the sum of all achieved BRS values and the theoretical maximum possible.

- **Example:** If three MCs were assessed, each with four cytokine indicators (maximum = 3.0), and the patient exhibited values of 0.5, 0.5, and 0.75, the total = 1.75. The GPR= 1.75 ÷ 3.0 = 0.583.

GPR, therefore, reflects the **global inflammatory load** contributing to cognitive decline.

### E) Global Cognitive Risk (GCR)

The GCR provided a **summative index of cognitive burden** across all MCs. It was calculated as the ratio between the total CRS achieved and the maximum achievable CRS.

- **Example:** If the maximum CRS = 200 and a patient’s score = 50, then GCR = 50 ÷ 200 = 25%.

GCR thus quantifies the **overall impact of systemic conditions on brain function**.

The IICM™ platform triangulates cytokine biomarker expression, standardized cognitive test performance, and systemic comorbidities to provide a root-cause analysis of the active promoters of cognitive decline. By combining immunologic and mental data, IICM™ enables the generation of individualized risk maps, supporting precision diagnostics and identifying potential therapeutic targets at preclinical and early clinical stages.

## Discussion

### Novelty and Rationale of the IICM™ Scoring Framework

The Inflammatory, Immunologic and Cognitive Mapping (IICM™) platform offers a new framework for quantifying and contextualizing the early promoters of cognitive decline. By developing unique condition-specific indices, the **Biomarker Risk Score (BRS)** and the **Cognitive Risk Score (CRS)**, and extending them into global indices of systemic risk, the **Global Patient Risk (GPR)** and **Global Cognitive Risk (GCR)**, the framework bridges three domains that have historically been assessed in isolation: biological signals, systemic comorbidities, and cognitive performance.

Traditional approaches to dementia detection typically rely either on molecular profiling (e.g., amyloid or tau biomarkers, genetic markers) or on neuropsychological testing once deficits are clinically apparent.^24-26^ These tools, while informative, are limited in two ways. First, they often identify pathology after neuronal damage has become established and less reversible. Second, they rarely integrate the contribution of modifiable systemic promoters such as vascular dysfunction, metabolic dysregulation, or systemic inflammation, factors now known to accelerate neurodegeneration years before clinical diagnosis.

The IICM™ scoring system directly addresses these limitations. By calculating the relative inflammatory burden (BRS) and linking it with measurable cognitive underperformance (CRS), the framework produces root-cause maps of decline that highlight actionable disease promoters. The GPR and GCR then provide a summative picture of an individual’s overall biological and functional risk, enabling clinicians to view dementia not as a binary diagnosis but as a dynamic, quantifiable continuum in a patient-specific manner. This approach offers a new perspective by linking systemic disease promoters with domain-specific cognitive impairment, and its clinical potential lies in its ability to reclassify patients at an earlier, more treatable stage.

### Preliminary Findings: Proof-of-Concept for Risk Stratification

Our pilot study provides early but compelling support for the validity and clinical utility of this framework.

- **Early Cognitive Decline (52%)**: Over half of the cohort demonstrated early-stage cognitive underperformance that conventional assessments would have likely missed. This finding underscores the sensitivity of IICM™’s cognitive mapping to detect subtle impairments across multiple domains. It also illustrates the added diagnostic value of integrating cognitive indices with systemic biomarker signals, as isolated cognitive testing often fails to capture underlying systemic contributions.
- **Biomarker–Cognition Correlations**: Strong associations between elevated cytokine expression and regional cognitive deficits further validate the construct of IICM™. These correlations provide mechanistic evidence that systemic inflammation is not merely an associated comorbidity but a measurable driver of functional decline. By linking these domains through structured risk scoring, IICM™ transforms diffuse biological and cognitive signals into a unified, clinically interpretable framework.
- **Pre-diabetes Detection (34%)**: More than one-third of participants were suspected to have pre-diabetes, a systemic condition strongly linked to neuroinflammation, vascular dysfunction, and accelerated cognitive decline. Importantly, pre-diabetes is both underdiagnosed and highly modifiable, making its early identification critical for preventive care. By capturing biomarker dysregulation attributable to metabolic pathways, the IICM™ platform highlights a silent but treatable promoter of decline.
- **The “Golden Hour” Cohort**: A subset of patients demonstrated elevated cytokine expression despite preserved cognitive performance. We define this stage as the *“golden hour”* of cognitive decline risk: a critical window in which systemic promoters of disease are already active but functional impairment has not yet emerged. Identifying patients within this transitional state may be pivotal, as it offers the most significant potential for preventive intervention before irreversible neuronal injury occurs. While preliminary, these findings suggest that IICM™ may provide a means of capturing disease risk during this otherwise invisible window, enabling clinicians to intervene at the point of maximal modifiability.

Together, these preliminary findings suggest that the BRS and CRS are capable of uncovering vulnerabilities that remain invisible to current diagnostic practices, while the global indices (GPR, GCR) provide a scalable method for quantifying overall disease burden.

### Clinical Implications: A New Tool for Early Intervention

The implications of this integrated approach for clinical practice are very promising.

#### Early Identification of Modifiable Risk

By detecting systemic promoters such as pre-diabetes, hypertension, or sleep apnea, IICM™ provides clinicians with an opportunity to intervene at a stage when cognitive decline may still be preventable or even reversible. Unlike amyloid or tau biomarkers, which confirm pathology but offer limited modifiability, the systemic risk factors revealed through BRS and CRS are directly actionable with lifestyle, pharmacological, and preventive strategies.

#### Personalized Risk Stratification

The dual scoring (BRS + CRS) allows clinicians to move beyond population-level risk factors and generate patient-specific risk maps. These maps highlight not only whether a patient is at risk, but why; whether due to metabolic, vascular, inflammatory, or combined pathways. This information can guide targeted interventions, from glycemic control to anti-inflammatory regimens or vascular protection, tailored to each patient’s underlying biology.

#### Improved Triage and Care Pathways

The global indices (GPR, GCR) offer simple, interpretable metrics that can be integrated into clinical workflows. Patients with elevated scores may be prioritized for specialist referral, advanced imaging, or eligibility screening for disease-modifying therapies such as anti-amyloid antibodies. Conversely, those with low global risk may be managed with preventive lifestyle modifications in primary care, reducing unnecessary costs and resource utilization.

#### Longitudinal Monitoring

Because the indices are quantitative and reproducible, they can be tracked over time, enabling clinicians to monitor disease progression and response to therapy. This opens the possibility of using IICM™ as both a diagnostic tool and a monitoring tool, applicable not only in clinics but also in the context of clinical trials evaluating preventive or disease-modifying therapies.

### Scientific Implications and Broader Significance

The IICM™ framework also contributes to the broader scientific understanding of dementia as a multifactorial disease process. By operationalizing systemic inflammation and metabolic dysfunction into structured risk scores, it provides a platform for testing hypotheses about causal pathways of decline. The biomarker–cognition correlations observed in this pilot support the emerging view that dementia is not solely a disease of the brain but one of systemic dysregulation with CNS manifestations.

Furthermore, the modular structure of the scoring system makes it adaptable to future biomarker discoveries. For instance, additional panels of cytokines, proteomic markers, or imaging-derived metrics could be incorporated into the BRS, while new digital cognitive tools could enhance CRS granularity. This flexibility ensures that IICM™ can evolve in parallel with advances in neuroscience and digital health.

### Future Directions and Potential Integration

Future studies will require larger, multi-center cohorts to validate the predictive accuracy and reproducibility of the IICM™ indices. Importantly, these studies should assess longitudinal outcomes to determine whether elevated BRS, CRS, GPR, and GCR predict conversion from preclinical stages to mild cognitive impairment or overt dementia.

Beyond validation, integration into primary care and community-based screening programs may represent the most significant opportunity for impact. Because the platform relies on peripheral blood biomarkers and digital cognitive testing, it is inherently scalable and does not require advanced neuroimaging or high-cost infrastructure. Linking IICM™ outputs to digital health platforms could further expand reach into underserved populations, helping to mitigate the persistent disparities in dementia diagnosis and care access.

The platform also has potential as a clinical trial endpoint. By quantifying mechanistic promoters of decline, IICM™ could serve as a surrogate marker for therapeutic response in trials targeting inflammation, metabolism, or vascular dysfunction. This would accelerate evaluation of preventive and disease-modifying strategies, while also enabling more precise selection of eligible participants.

Finally, this preprint represents the first stage of our research program. We are actively expanding the research and advancing the methodology to increase predictive robustness and clinical applicability. The full dataset, including extended analyses and validation, will be submitted for peer-reviewed publication in the near future. By sharing these early findings now, we aim to stimulate dialogue, invite collaboration, and accelerate progress toward a scalable, precision-based approach for the early identification of cognitive decline and provide potential opportunities for intervention.

## Conclusion

This pilot study demonstrates the feasibility and potential of the IICM™ platform to identify early, systemic promoters of cognitive decline using a novel, integrative risk scoring framework. Preliminary results, highlighting pre-diabetes detection, early cognitive underperformance, and biomarker–cognition correlations, support the platform’s validity and clinical relevance. By transforming complex multidimensional data into actionable risk maps, IICM™ offers a promising path toward earlier diagnosis, targeted prevention, and precision management of dementia.

## Data Availability

All data produced in the present study are available upon reasonable request to the authors

http://www.ascento-cda.com

