## Appendix 1 for "Early Identification of Disease Promoters of Cognitive Decline Using Inflammatory, Immunologic and Cognitive Mapping (IICM™)"

### Data analysis

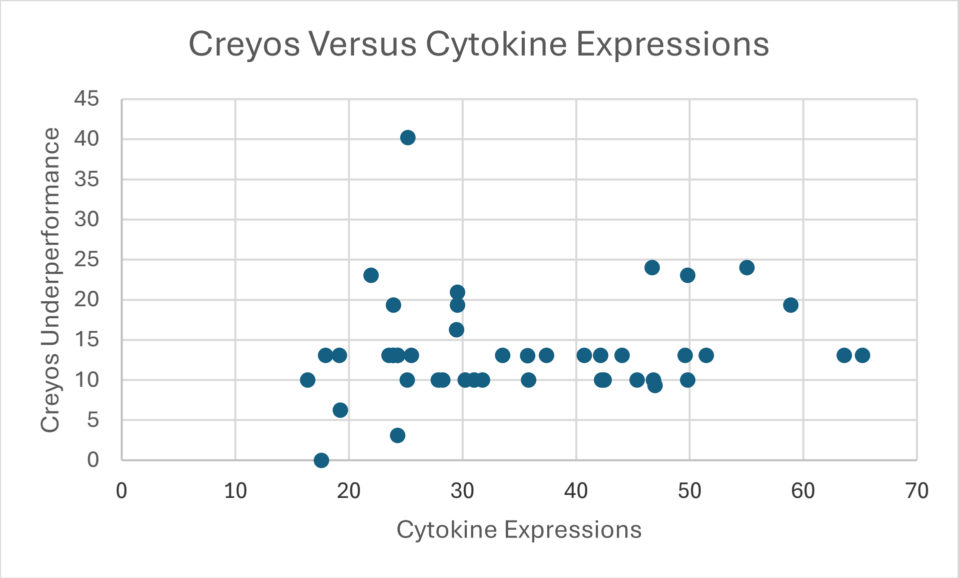


Fig. 1: The expression of Cytokines versus Creyos grades:

The higher the number of Cytokine expressions found, the high the Cytokine Expression score. This constitutes GPR (Global Patient Risk)

The more a patient underperformed within Creyos, the **higher** the Creyos score, which constitutes the GSR (Global Cognitive Risk).

We found that 28 out 45 participants (63%) had cognitive decline that was expressed within co-morbidities.

This is based on a threshold of 15% decline, as measured by Cognitive Patient Risk.

##### Pre-diabetes Detection

We defined a marker of 3 over-expressed Cytokines, relevant to diabetes as predictors of diabetes or pre-diabetes.

We assumed that 4 or 5 over-expressed Cytokines would already have been detected by the medical teams.

In this study we did not rely on self-declaration reg. the actual diagnosis of diabetes.

Distribution of patients:

| Number of over-expressed cytokines | 5 | 4 | 3 | 2 | 1 | Total |
|---|---|---|---|---|---|---|
| Number of patients | 3 | 8 | 15 | 14 | 5 | 45 |

The table indicates that 11 patients had 4 or 5 over-expressed Cytokines. We consider them advanced enough in the medical prognosis to be identifies by other means.

For 15 patients, we found 3 over-expressed Cytokines, which we consider high-risk for pre-diabetes or un-diagnosed diabetes.

Within 19 patients, 1 or 2 Cytokines were over-expressed, which we consider too low to justify flagging as at-risk patients reg. Diabetes
